# Tangible advantages of multi-stage brain motion compensation for PET imaging demonstrated in multiple studies

**DOI:** 10.1101/2025.09.18.25336066

**Authors:** Nikos Efthimiou, Jessie Fanglu Fu, Mehrbod Mohammadian, Chieh-En J. Tseng, Michele Scipioni, Iris Y. Zhou, Nicole R. Zürcher, Joseph B. Mandeville, Julie C. Price, Marco L. Loggia, Ciprian Catana

## Abstract

Head motion is a persistent challenge in positron emission tomography (PET) brain imaging, reducing quantitative accuracy, degrading time-activity curves (TACs), and complicating kinetic modeling. We evaluated a fully automated, multi-stage motion compensation framework, COMBRA (Correction of Motion and BRain Alignment), across more than one hundred scans and four independent PET studies using different radiotracers (^18^F-MK-6240, ^11^C-Raclopride, ^11^C-PBR28, and ^11^C-Martinostat). COMBRA implements a staged registration and reconstruction pipeline, enabling both inter-frame realignment and intra-frame motion-gated correction. It operates in a PET-driven mode but can incorporate magnetic resonance imaging (MRI)-based motion tracking when available. Across dynamic datasets, COMBRA improved model fitting precision, as reflected by reduced standard error in distribution volume ratio estimates using MRTM2 modeling, and yielded more consistent test– retest outcomes. In static cohort studies, the framework enhanced regional standardized uptake value ratios, improved image contrast, and mitigated motion-induced bias. Compared to frame-to-frame realignment and MRI-guided approaches, PET-only COMBRA demonstrated equivalent or superior performance, with misalignments reduced to sub-voxel levels. Importantly, its static reconstructions provided better signal-to-noise properties and more robust quantification in high-motion cases. At the population level, COMBRA increased statistical power, suggesting that smaller sample sizes may be sufficient for future studies. Collectively, these results demonstrate that COMBRA offers a scalable, tracer-independent solution for motion compensation in brain PET imaging. By improving accuracy, reliability, and sensitivity, this framework has the potential to strengthen both mechanistic research studies and clinical investigations where subtle group differences are critical.

## 1. Introduction

Head motion remains a challenge in positron emission tomography (PET), introducing blurriness, degrading quantitative accuracy, and hindering image analysis and kinetic modeling [1–8]. Motion causes misregistration between emission data and the anatomical image used for attenuation correction (*μ*-map), as well as the regions of interest (ROIs) used for analysis and voxel-level parametric maps. This impairs quantification, increases TAC noise, reduces curve-fitting precision, and raises residuals, complicating modeling. ROI misalignment further exacerbates analysis challenges.

Multiple strategies have been developed to reduce or compensate for motion effects, categorized by when motion information is used: pre-, during, or post-reconstruction [5,6,9,10]. All approaches require motion estimation, which can be derived from external markers [11–13], devices [14], simultaneously acquired magnetic resonance imaging (MRI) data, or directly from PET [15–23].

Numerous automatic inter- and intra-frame motion correction (MoCo) pipelines have been proposed [24], many relying on good anatomical-functional correspondence [25]. However, PET radiotracers often show poor spatial correlation with anatomy—especially in long dynamic acquisitions with high-affinity tracers [26]. For instance, early PET frames often reflect blood flow and align better with MR, while later frames reflect tracer binding. [^18^F]-MK6240, for example, binds to tau aggregates localized in the temporal cortex in early Alzheimer’s disease (AD). In contrast, FDG reflects whole-brain glucose metabolism and aligns more consistently with anatomical structure. Recent machine learning (ML)-based MoCo methods show promise [25] but require large training datasets, which are difficult to obtain for many tracers.

Despite progress, a need remains for adaptable, vendor- and tracer-independent solutions to ensure accurate quantification across PET studies. To address this, we developed a fully automated framework: COrrection of Motion and BRain Alignment (COMBRA). It provides a data-driven solution for inter-frame activity realignment and parallelized intra-frame multi-gated motion-compensated reconstruction. This supports static reconstructions and enables user-directed frame adjustments in dynamic scans when COMBRA’s automatic settings are insufficient. COMBRA also supports optional MRI-guided motion estimation.

In this paper, we illustrate the advantages and discuss the limitations of the proposed method by using data from four studies conducted at our institution. We chose to assess the impact of the proposed framework while adhering to each study’s original methodology and endpoint. Additionally, we ensured that the reconstruction and post-reconstruction filtering settings were closely aligned with those used on the scanner to minimize discrepancies. The studies fall into two groups: dynamic studies, focused on kinetic modeling using ^18^F-MK-6240 (MK) and ^11^C-Raclopride (RAC), and static studies, utilizing semiquantitative analysis using ^11^C-PBR28 (PBR28) and ^11^C-Martinostat (MSTAT). The framework’s impact was assessed based on diverse endpoints, including cohort separation into different motion subgroups, the accuracy of kinetic modeling, and comparisons with existing MoCo techniques such as frame-to-frame PET realignment and MRI-guided methods.

## 2. Materials and Methods

### 2.1. COMBRA Framework

COMBRA employs staged, multi-resolution registrations for dynamic and static motion-corrected PET reconstructions, summarized in Fig. 1. The inputs consist of the list-mode PET data, the μ-map volume generated from an anatomical imaging modality and, optionally, a skull-stripped MRI image as extracted from FreeSurfer (http://surfer.nmr.mgh.harvard.edu). The workflow begins with the generation of dynamic frames from the PET list-mode data.

**Figure 1.**
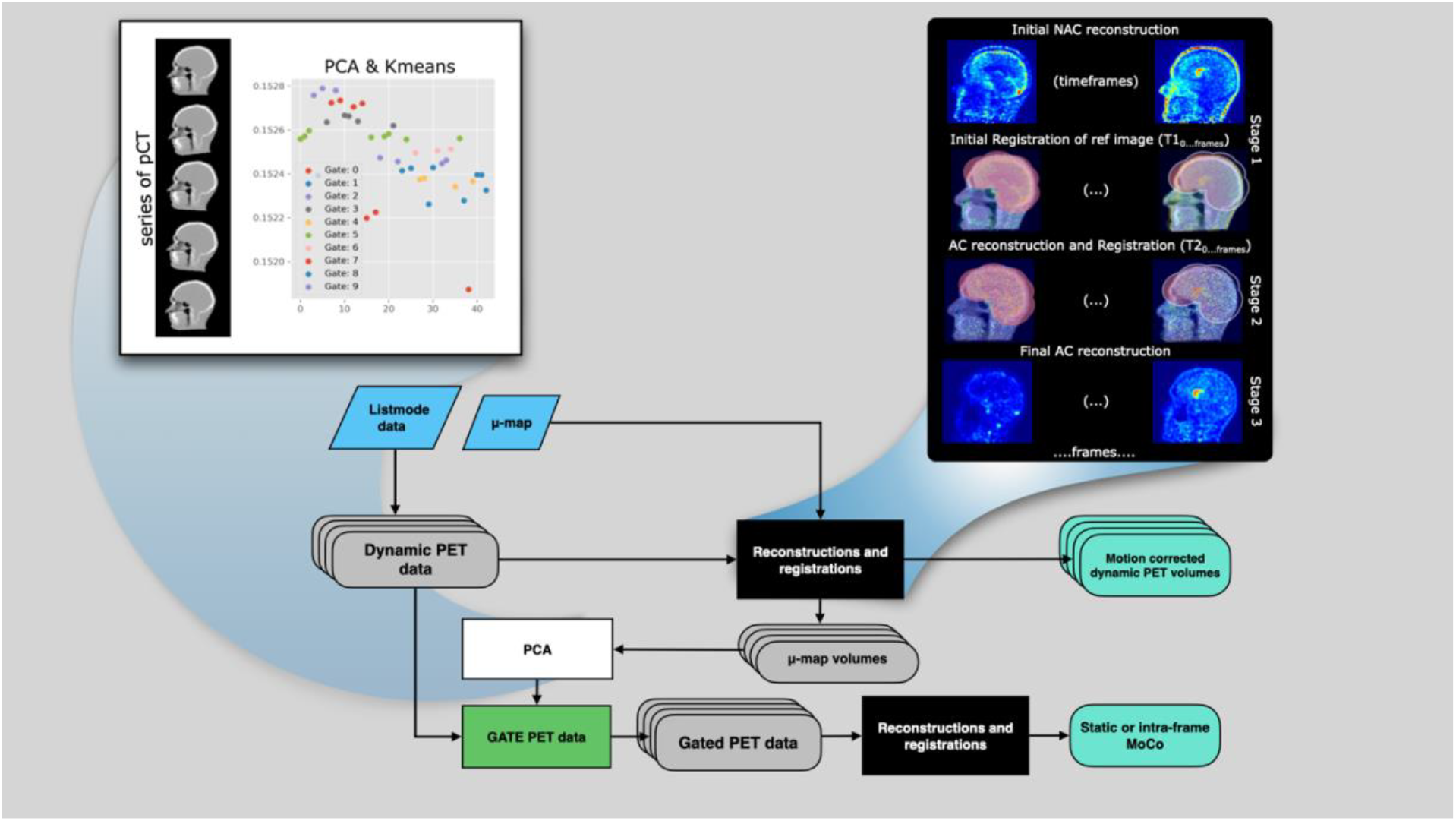
The COMBRA schematic flowchart for dynamic and static motion compensating image reconstruction. The input to COMBRA is the listmode data and the μ-map (pCT). The black box details the Reconstruction and Registration (R’nR) loop. The white box details the process of getting the motion surrogate signal with PCA over the anatomical images and Kmeans clustering, used for gating the PET data.

In the MoCo COMBRA pipeline, dynamic PET data are reconstructed, and anatomical volumes (typically μ-maps) are registered with the PET volumes in stages using a reconstruction and registration (R’nR) loop with an increased number of iterations in each stage. The R’nR starts with low-noise non-attenuation-corrected (NAC) reconstructions of the PET frames and registration of the original μ-map on each PET frame (stage 1). In the next stage, attenuation corrected (AC) reconstructions are performed, using a registered μ-map for each PET frame, and a second, refined registration (stage 2). Finally, the process ends with a last round of reconstructions and post-reconstruction filtering.

PET frames can be generated based on predefined time intervals or with a recorded counts threshold. In dynamic studies, either option is available. However, if static images or intra-frame MoCo are the endpoints, then low-count frames with 2.5×10^6^ events each are automatically generated and used.

After the first execution of the MoCo pipeline, principal component analysis (PCA) on the series of resulting μ-maps is performed and the eigenvalues of the first component are extracted. This component holds information on the covariance between the frames, typically encapsulating the slowest movements. The eigenvalues are then clustered to form multiple PET gates, each with a minimum of 7.5×10^6^ true events (see supplemental material A). Finally, the gated PET sinograms are reconstructed through a second pass of the MoCo pipeline. Reconstructed gated PET images are weighted, merged, and corrected for decay and frame length. In all cases, Gaussian filtering is applied as needed.

COMBRA is independent of the modality used to acquire the anatomical data; it can operate with data from either CT or MRI. We used T1-weighted MRI and derived pseudoCT (pCT) images [27]. The pCTs have shown better robustness in cases where PET tracer uptake poorly correlates with the T1 contrast. Additionally, COMBRA has an MRI-guided motion estimation “mode” that allows pCTs or T1-weighted images to be registered to MRI echo planar imaging (EPI) volumes when acquired during PET scans (see supplemental material B).

Image registration was performed using Greedy (v1.3) [28], a software implementing stochastic quasi-Newton optimization with Limited-Memory L-BFGS via the VNL library. Normalized Mutual Information (NMI) was the cost function (60 bins, linear interpolation) across three resolution levels (x4, x2, x1), smoothed with 2, 1, and 0 mm sigmas, respectively. Random jitter (0.5 voxels) was applied at each resolution.

PET signal activity and anatomical images were automatically cropped around the head and neck using SciPy’s find_objects with a relaxed bounding box to optimize efficiency and metric accuracy. A “bubble”-head mask, derived from FreeSurfer skull-stripped brains [29,30] and dilated with a spherical kernel, constrained registration to the brain area (illustrated with pink color in Fig. 1). This mask was applied to the fixed PET image, restricting cost function calculations and mitigating face and neck interference.

Registration robustness was enhanced through a majority voting scheme based on three independent normalized mutual information (NMI) calculations: Greedy’s native implementation, a Python-based implementation using scikit-learn [31], and SimpleITK’s function. The NMI was also evaluated pre-registration; if superior, registration was aborted, and the anatomical image from the closest time frame was substituted. The methods described were consistently applied in the MRI-guided mode.

### 2.2. Datasets

Subjects were retrospectively selected from studies previously conducted at our institution on a Siemens Biograph mMR (Siemens Healthineers, Erlangen, Germany), an integrated PET/MRI scanner. All participants provided written informed consent, and the studies were approved by the institutional review board. Across all studies we used data from 33 healthy controls, 3 patients with Alzheimer disease (AD) and 54 patients with chronic low back pain.

### 2.3. Dynamic studies

#### 2.3.1. MK study

Dynamic PET data were acquired for 120 minutes post-injection with a 15-minute break at 65 minutes. The first acquisition was broken into 29 dynamic frames, starting at injection, with durations of 6×10-, 6×20-, 2×30-, 2×60-, 2×120-, and 11×300 seconds. The second acquisition consisted of 8×300-second frames.

The ROIs analyzed were the entorhinal, inferior temporal, parahippocampal, amygdala, and hippocampus regions. These regions typically show early signs of tau burden across the AD spectrum and are particularly susceptible to head motion bias due to their small sizes. The multilinear reference tissue model (MRTM2) was employed to estimate distribution volume ratio (DVR) values, with t*=30min, k2’=0.04, and cerebellar gray matter as the reference region [33,34].

The DVR standard error (*DVR*_*SE*_) was used to assess the impact of MoCo on MRTM2’s fit quality. Additionally, the percentage differences between test-retest cases were examined using the different MoCo methods (supplemental material E).

#### 2.3.2. RAC study

This study aimed to illustrate variations in ^11^C-Raclopride (a selective D2/D3 dopamine receptor antagonist) uptake binding potential resulting from a behavioral task involving monetary rewards, which stimulates the basal ganglia.

Participants received a bolus injection, followed by a dynamic PET data acquisition for 90 minutes. The initial study utilized 47 time frames with incrementally longer durations (22×60 -, 12×120 -, 8 × 180 -, 5 × 240 seconds).

The ROIs analyzed were the putamen, caudate, nucleus accumbens, and thalamus. The full reference tissue model (fFTRM) was employed to estimate regional binding potential measures and the quality of the model fits were evaluated [33]. The inverse residual coefficient of variation (IRCV) was calculated as the mean TAC value divided by the standard deviation (SD) of the residuals. The IRCV quantifies relative variability, indicating the extent of mean signal change in relation to its residual SD. This metric can be interpreted as a temporal, model-dependent signal-to-noise ratio (tSNR).

The MRI-based MoCo used for comparison was applied post-PET image reconstruction using the vendor supplied OSEM algorithm (21 subsets and 3 iterations). For each PET frame, a time-averaged MRI volume was created from all EPI volumes that were acquired during the specific PET frame interval [33]. These EPI volumes were coregistered using AFNI [36], and the 6-dimensional rigid-body motion parameters were applied to the PET data for each time frame.

The simultaneously acquired slice-selective EPI had a 3 mm isotropic resolution (TR/TE = 3000/30 with 42 slices covering the full brain), excluding the 20-minute task period (TR/RE = 2000/20 with 34 slices). The images were reconstructed with 2-fold acceleration (GRAPPA) in the phase direction.

### 2.4. Static studies

In both static studies, the imaging protocols involved co-registering T1-weighted MRI volumes with PET volumes reconstructed from the data acquired during the 60-to 90-minute post-injection period. The standard uptake values (SUV) (g/mL) maps were generated by normalizing PET uptake to injected dose and body weight.

SUV maps were intensity-normalized by the mean SUV extracted from the whole brain, resulting in SUV ratio (SUVR) maps. Image analyses were performed using FSL (FMRIB SoftwareLibrary, http://www.fmrib.ox.ac.uk/fsl), AFNI (Analysis of Functional NeuroImages, (http://afni.nimh.nih.gov/afni) and FreeSurfer. Processing was performed in subject space for MSAT and in Montreal Neurological Institute (MNI) space (MNI152) FOR PBR28.

#### 2.4.1. MSTAT study

This study explored the involvement of epigenetics in socio-emotional behavior (SEB) and sex differences in epigenetics by employing ^11^C-Martinostat PET to evaluate in vivo histone deacetylases (HDACs) in young, healthy adults.

Based on previous findings, the amygdala and hippocampus were the designated ROIs to assess sex differences in HDACs [37,38]. ^11^C-Martinostat exhibits a strong visual correspondence with contrast differences between white and gray matter in MPRAGE T1-weighted images. This characteristic makes it an excellent candidate for comparing COMBRA MoCo using the anatomically rich MPRAGE images against the pCT maps.

#### 2.4.2. PBR28 study

The IGNITE was a clinical trial to evaluate the potential anti-neuroinflammatory effect of minocycline in chronic low back pain. The 18 kDa translocator protein (TSPO) is a potential neuroinflammation indicator. Research, using dedicated brain PET/MRI, has shown elevated TSPO levels in chronic low back pain patients, particularly in the thalamus region [39,40]. Subjects underwent imaging using ^11^C-PBR28, a radioligand that binds to TSPO. This study has the largest cohort in this paper, allowing us to separate the subjects into three groups (low-, mid-, and high-movers) based on the aggregated motion (see supplemental B).

## 3. Results

### 3.1. Benefits to kinetic modeling

#### 3.1.1. Modeling with MRTM2

As shown in Fig. 2.A, COMBRA demonstrated significantly reduced mean DVR and narrower 95% confidence intervals compared to NoMoCo and an in-house developed frame-to-frame PET alignment (F2F) across most brain regions, with improvements most pronounced in AD subjects due to their higher head motion. In the entorhinal cortex, COMBRA reduced DVR by 51% compared to NoMoCo, outperforming F2F’s 27% reduction.

**Figure 2.**
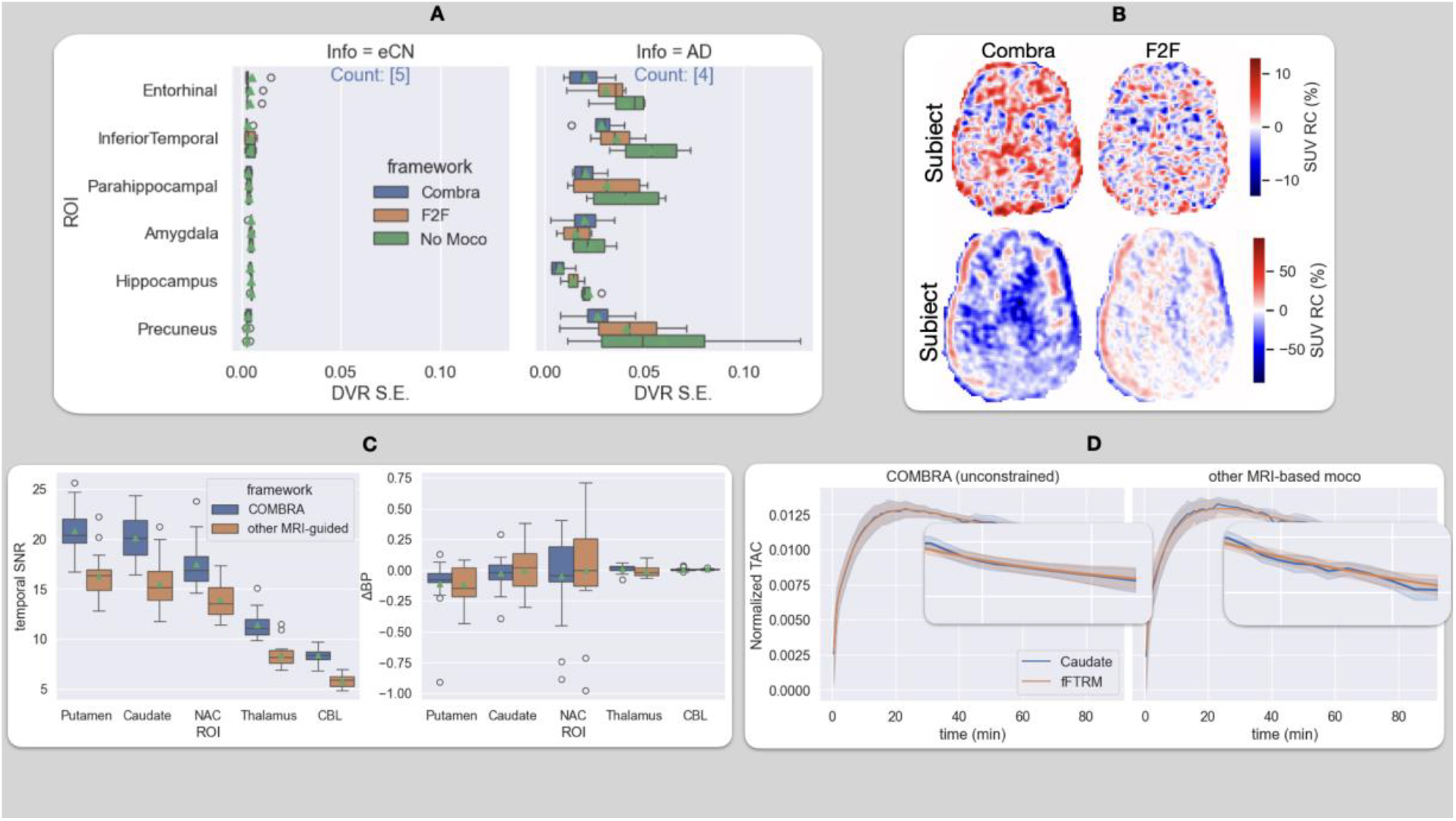
A.Distribution of DVR_SE_values for all subjects in ^18^F-MK-6240. B. Relative change images for a low- and high-moving subject (Subjects A and B, respectively) between the noMoCo and the two MoCo pipelines. C.(left) Improved temporal SNR in 11C-Raclopride TACs. (right) Distribution of ΔBPnd values for different regions of interest. D. Comparison between the average TACs of the PET-only COMBRA with unconstrained PET frames and in-house developed MRI-guided post-reconstruction activity realignment and time-based frames.

Similarly, COMBRA achieved a 64% reduction in the hippocampus compared to F2F’s 35%. However, F2F performed slightly better in the amygdala, reducing errors by 34% compared to COMBRA’s 15%. Healthy elderly controls (eCN) showed minimal DVR*SE*across all methods.

Masked relative change (RC) images revealed that COMBRA exhibited the largest differences from NoMoCo for both low-motion eCN and high-motion AD subjects (Fig. 2.B). MoCo with COMBRA increased apparent uptake in low-motion cases, while in high-motion cases, it was reduced.

The masked RC images for COMBRA and F2F from the NoMoCo images for Subject A (eCN, low mover) and Subject B (AD, high mover) are shown in Fig. 2.B. In both cases, COMBRA shows the largest differences from NoMoCo. Interestingly, in the eCN low-mover, the difference is mostly positive, indicating that MoCo increased the apparent uptake. However, in the AD high-mover, the opposite was observed.

#### 3.1.2. Modelling with fFTRM

As shown in Fig 2.C, COMBRA improved tSNR by approximately 30% across all ROIs compared to an in-house developed post-reconstruction MoCo method. Despite no significant differences in mean BPnd between the methods, COMBRA achieved a substantial reduction in interquartile range (IQR) across subjects in all regions, reflecting enhanced fitting precision.

The average TAC and fFTRM model for the caudate, shown in Fig. 2.D, highlight improvements in noise reduction (blue curves) and precision (orange curves). These enhancements are particularly notable around the peak uptake (20 minutes post-injection) and during the 40-60-minute interval, where significant head motion occurs.

COMBRA’s event-based mode produced smoother TACs beyond 60 minutes but with fewer data points, leading to an increased CI. Additionally, the time-based PET frames introduced bias in the TAC tails due to the short half-life of ^11^C, further emphasizing the advantages of COMBRA’s event-based framing.

### 3.2. Benefits in Static PET

Fig. 3.A illustrates aggregated head motion in the ^11^C-Martinostat cohort, showing minimal motion (below voxel size) for most participants, except for a single frame from Subject 7, who moved approximately 4 mm in the final PET frame. Sagittal slices of reconstructed PET frames overlaid on anatomical images, with and without registration.

**Figure 3.**
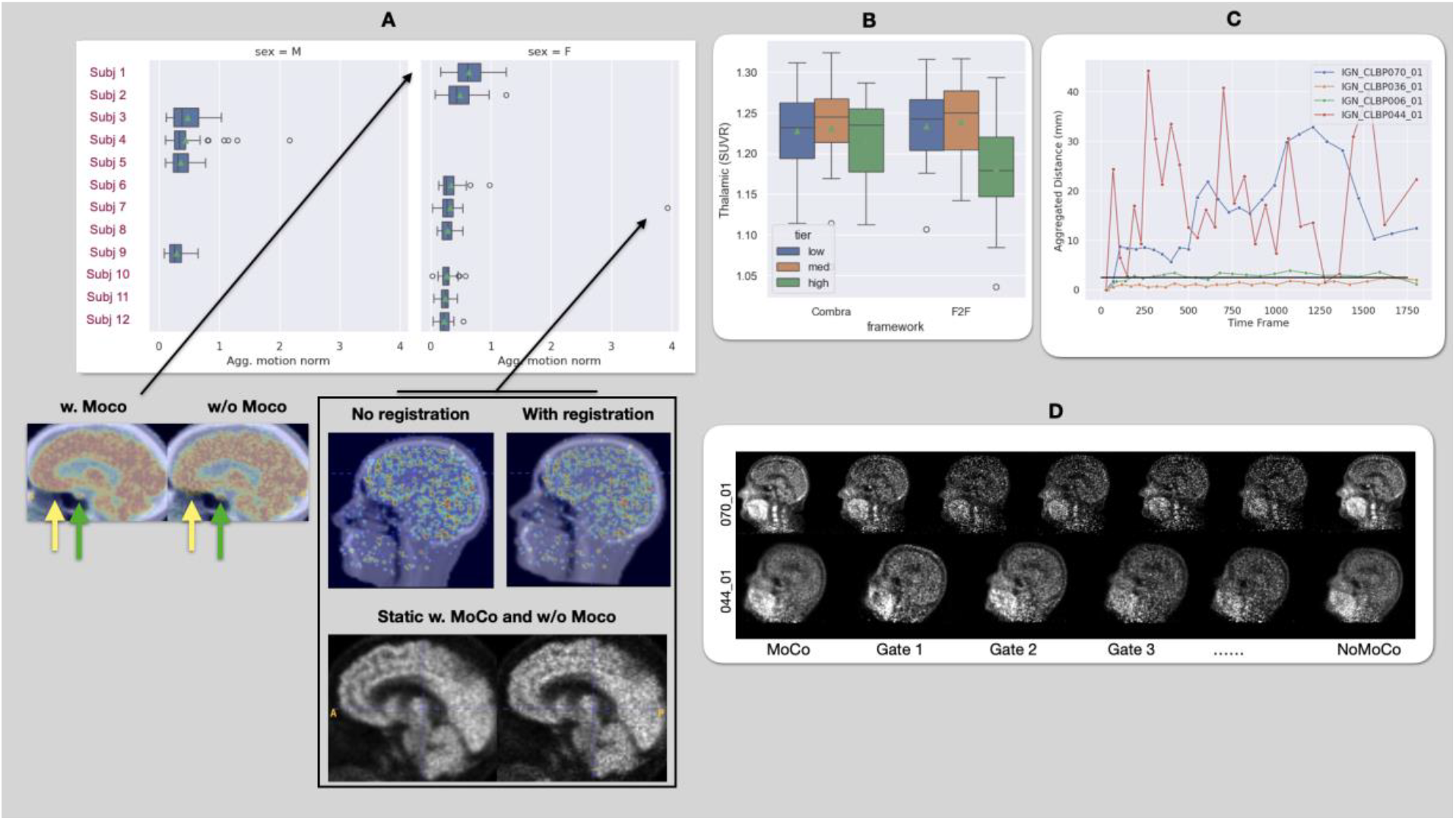
A.MSTAT subjects sorted by aggregated motion across the dynamic scan. We also show static images with and without MoCo for the subject with the highest average motion and, in the box, the subject with the highest motion in a single dynamic frame superimposed on the attenuation map: single frame (top) and whole acquisition (bottom). Yellow arrows point to misalignment between PET and anatomical images that impaired local quantification, while green arrows indicate improvements in the pituitary gland. B. PBR28: at the group level, after MoCo, SUVR values of high-movers approached those of low- and mid-movers. C. Aggregated motion for a low-mover (036), mid-mover (006), and two high-movers (070, 036). The horizontal black line indicates voxel size. D. Slices shown for MoCo, NoMoCo, and each gate for high-motion subjects from C.

For Subject 1, who exhibited the highest average motion, static PET images after MoCo show improved anatomical alignment. The pituitary gland (green arrows) and thalamus are more distinctly shaped, and the brain aligns accurately with the anatomical map (yellow arrows). These results highlight MoCo’s ability to correct motion artifacts and enhance image quality in static PET studies.

In the PBR28 study, the subjects were categorized into high-, mid-, and low-movers based on aggregated motion levels. Group-level analysis of thalamic SUVR revealed minimal MoCo impact for low-and mid-movers, whose motion was below or near voxel size. However, high-movers showed an increase in mean SUVR after MoCo, although its effectiveness was limited by the small number of collected events (Fig. 3.B). The described analysis highlights the limitations of MoCo when the number of collected events is insufficient.

High-movers, who exhibited an average aggregated motion of approximately 7.5 mm per gate compared to 0.5 mm for low-and mid-movers, showed wider eigenvalue spreads across frames, indicating greater motion variability (Fig. 3.C). This variability challenges MoCo’s effectiveness, especially when only four gates are assigned, as it is limited by the collected events (Subject 044_01).

However, increasing the number of gates improves image quality (Subject 070_01), a high-mover with sufficient events to allow an additional gate (Fig. 3.D). This subject’s motion-corrected image showed reduced motion artifacts compared to 044_01, emphasizing the importance of adequate event collection for effective MoCo in PET.

## 4. Discussion

Motion-induced artifacts remain a significant confounding factor in brain PET quantification, particularly in dynamic studies and in imaging of neurodegenerative disorders [8,41,42]. This study demonstrates the advantages of COMBRA, a parallelized and parametrizable MoCo framework designed to improve accuracy and precision in quantitative PET imaging. By integrating COMBRA into multiple studies at our institution, we demonstrated its utility across diverse datasets and imaging protocols.

COMBRA’s ability to batch-process subjects makes it particularly advantageous for large retrospective studies, where head motion is a common issue. This capability not only enhances statistical power but could also reduce the required sample size. Additionally, its event-based dynamic framing approach ensures that each frame contains sufficient counts for accurate scatter correction and minimizes noise-related bias, as demonstrated in the RAC study.

In the MK study, DVR_*SE*_was reduced by over 35%, with the most pronounced improvements observed in high-motion AD subjects compared to low-motion eCN controls. For example, in the inferior temporal lobe, interquartile ranges of DVR_*SE*_were reduced by 95%, highlighting COMBRA’s ability to mitigate motion-induced variability. This reduction in DVR variability can potentially improve the accuracy and sensitivity for quantifying tau burden and tracking longitudinal tau accumulation in AD.

Consistent with findings in the literature, MoCo does not alter the average MK DVR values at the population level (data not shown) [8,39]. This is because global fits across all frames can mitigate the impact of motion in individual frames, resulting in nearly identical average DVR values. However, MoCo notably enhances the precision of parametric models used in tracer kinetic analysis.

Interestingly, MoCo increased DVR SD at the group level for AD subjects by 26.8%, 7.94%, 55.3%, 43.5%, and 132.4% in the entorhinal, inferior temporal, amygdala, parahippocampus, and hippocampus regions, respectively. Despite the small cohort (4 AD datasets), we hypothesize that this likely reflects more accurate tracking of individual disease variability—a critical factor for understanding pathology and therapeutic evaluation. On the other hand, in the eCN, we observed a small reduction in the SD of the DVR.

In static studies using ^11^C-Martinostat, where head motion was minimal (below voxel size), MoCo showed limited group-level benefits but improved visual quality and alignment with anatomical images.

For high-motion subjects with aggregated motion exceeding 30 mm, benefits were constrained by the limited number of detected events per gate. Data collected from high movers often allowed only four gates—similar to low-motion subjects—resulting in up to 7.5 mm of residual motion per gate. This limitation highlights the need for advanced reconstruction techniques that reduce noise-induced bias and enable finer gating.

COMBRA outperformed other pipelines, such as the F2F and MRI-assisted MoCos implemented in-house. For high-movers, COMBRA improved skull delineation, reduced noise in the thalamus, and enhanced clarity in small structures such as the pituitary gland. While F2F averaged attenuation maps across frames to mitigate motion effects, it failed to effectively address intra-frame variability. Similarly, MRI-assisted post-reconstruction realignment methods showed limited improvements due to their reliance on time-based framing and reduced quantification.

Despite its advantages, COMBRA’s reliance on traditional reconstruction methods limits its ability to fully exploit advanced regularized algorithms [45] that could further reduce noise-induced bias and improve the contrast-to-noise ratio. Incorporating these methods could enable finer gating for high-movers and further enhance image quality. Additionally, early PET frames acquired during injection often lack sufficient counts for reliable head tracking. However, if necessary, MRI guidance could assist during these stages in integrated PET/MRI scanners. Future work will focus on integrating advanced reconstruction techniques [43,44] into COMBRA and optimizing gating strategies for high-movers to maximize its potential in both research and clinical settings.

## 5. Conclusion

COMBRA represents a robust solution for addressing motion artifacts in PET imaging. It improves precision at both individual and group levels while maintaining compatibility with diverse datasets and protocols. By enhancing parametric modeling accuracy and reducing variability caused by head motion, COMBRA is particularly valuable for neuroimaging studies of AD and other conditions where subtle changes are critical for understanding disease progression and evaluating interventions. Its event-based dynamic framing approach further underscores its potential as a versatile tool for improving PET quantification in challenging scenarios involving short-lived isotopes or low-injected dosages.

## Data Availability

All data produced in the present study are available upon reasonable request to the authors

## Acknowlegdements

Funding for this work was supported by NIH grant R01MH128425-01 (to NRZ). [^11^C]Martinostat PET-MRI data used in the preparation of this manuscript are available from the National Institute of Mental Health (NIMH) Data Archive (NDA). NDA is a collaborative informatics system created by the National Institutes of Health to provide a national resource to support and accelerate research in mental health. Dataset identifier: 10.15154/es4w-zn36. This manuscript reflects the views of the authors and may not reflect the opinions or views of the NIH.

