## supplements for "Tangible advantages of multi-stage brain motion compensation for PET imaging demonstrated in multiple studies"

### SUPPLEMENTARY MATERIAL

#### A. BIAS IN SHORT DYNAMIC FRAMES

The origin of the bias in the late part of the TACs is illustrated in Fig S1. When the number of net trues (prompts - randoms) per frame drops below a certain threshold (≈ 7.5 × $10^6$ events - horizontal line), the scatter fraction (S.F.) is overestimated, which leads to the steeper TAC slopes observed in Fig. **2**.D. Although increasing the frame duration (from 60 to 120 s) partially offsets the $^{11}C$ decay, after 300 seconds (frames with durations of 180 s and 240 s), the S.F. is overestimated. The S.F. of the unconstrained approach has a slight downward slope due to random scatter events that cannot be correctly addressed by single scatter estimation.

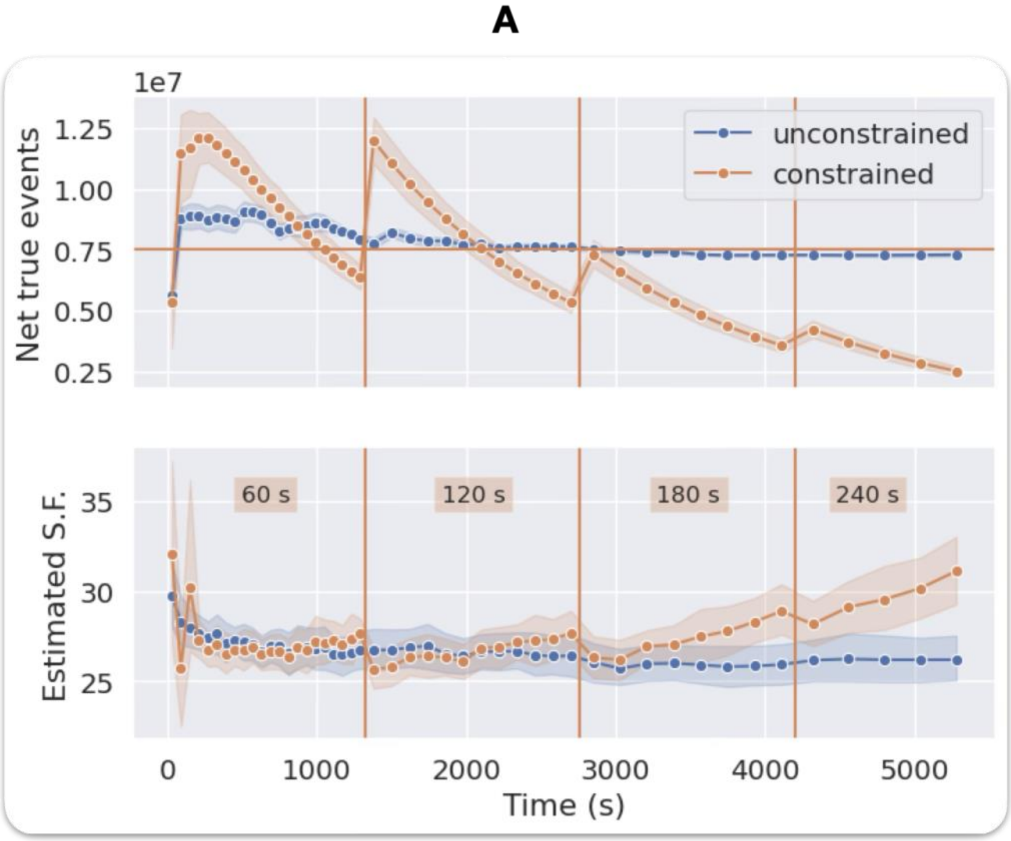


**Figure S1** Impact of the number of detected net true events on the accuracy of estimated scatter fraction, averaged over the study's cohort. When the events dropped below the threshold indicated by the horizontal line, bias was introduced in the scatter fraction estimation. Event-based (unconstrained) frames provide unbiased TACs.

#### B. DISCREPANCY BETWEEN PET-ONLY AND MRI-GUIDED HEAD TRACKING IN COMBRA

Low-resolution echo-planar images (EPI) were acquired during the first MK subject scans (TE=12 ms, TR=4800 ms, slice thickness=7.5 mm) and corrected for distortions using Freesurfer. These EPI volumes were used for head tracking in MRI-guided COMBRA to validate the PET-only approach.

Figure S3 compares aggregated motion between EPI-based and PET-only COMBRA for healthy controls and AD subjects. The average motion vector difference per PET frame was below 0.5 mm for most of the acquisition, well below the voxel size (2.1 × 2.1 × 2 $mm^3$). The mean difference increased toward the end due to delays between PET radioligand injection and MRI start times, leaving late PET frames uncovered by EPIs acquisitions, which were replaced with prior EPI volumes. Additionally, short EPI acquisitions provided limited coverage of longer PET frames. Reliance on MRI images can sometimes be inconsistent due to deviations from planned protocols.

**A**

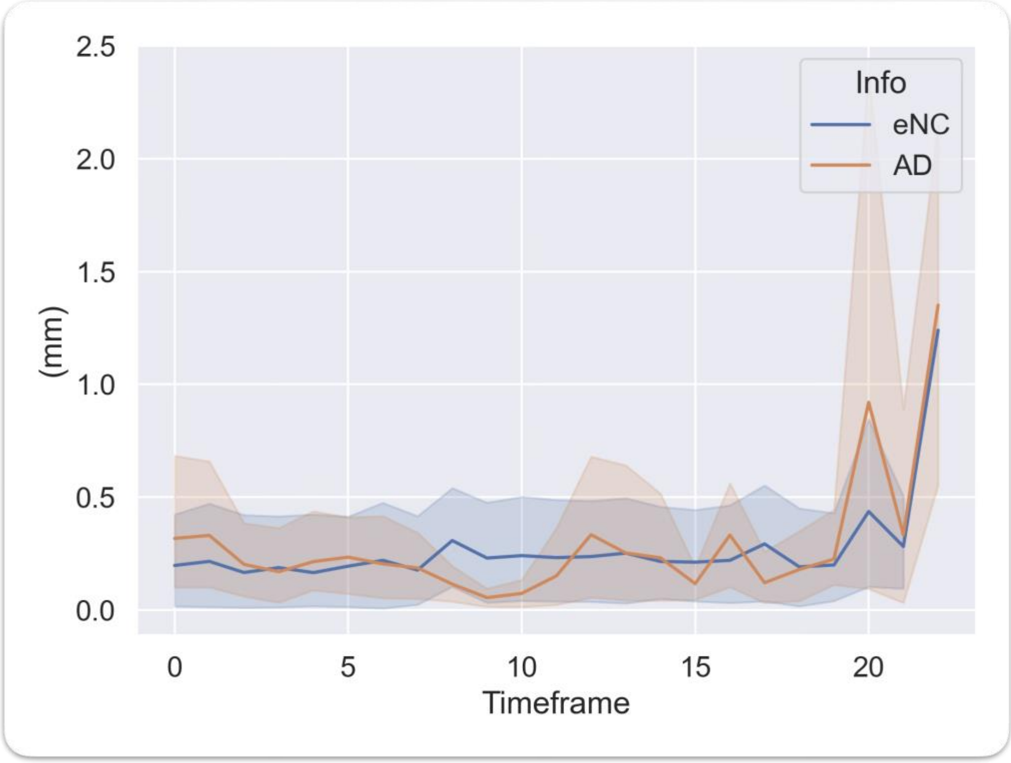


**Figure S2** Discrepancy between the MR-guided and PET-only COMBRA, as the difference between the estimated aggregated motion for the first 40 min of the MK study. (eCN) healthy elderly controls, (AD) Alzheimer's disease subjects.

#### C. MOTION EVALUATION

An aggregated motion assessment technique was implemented as previously described [1, 2] to quantify each subject's displacement index. A sphere with 512 equidistant points was algebraically simulated using the golden-angle Fibonacci algorithm. Transformations were applied to this sphere by inverting the estimated transformation parameters for each dynamic PET frame.

The aggregated motion was determined as the mean Euclidean distance between the original and transformed point positions across timeframes. Subjects were classified into "high," "medium," or "low" mover groups based on their maximum detected displacement.

#### D. RECONSTRUCTION QUANTIFICATION

To validate the quantification of static and gated images reconstructed with COMBRA, a stationary NEMA-type body phantom containing two rings of spheres (diameters: 10, 13, 17, 22, 28, and 37 mm) with an 8:1 sphere-to-background activity ratio was used.

Quantification was assessed by splitting the stationary phantom data into one, four, and ten gates using the PCA/K-means method. Each gate was reconstructed and compared to the static reconstruction of the full dataset. The mean activity concentrations for static, single-, four-, and ten-gated images were 15827.063, 14766.662, 15134.448, and 15286.673 kBq/mL, respectively, showing an average absolute quantification difference of 4% (Fig. S2). Corresponding contrast recovery ratios were 8%, 7.8%, 7.83%, and 7.6%. The observed contrast loss is likely due to the use of inverse transformations instead of adjoint operations.

**A**

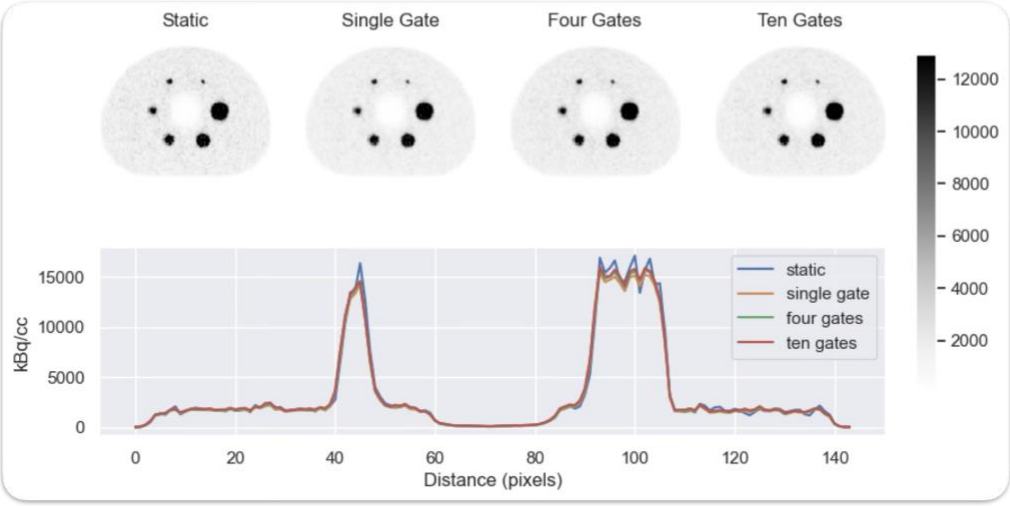


***Figure S2*** *A. Axial slices, centered on one ring of spheres of the phantom, reconstructed (left) with typical static reconstruction and then after gating the data into one, four, and ten gates. Most subjects in this study used three to five gates.*

#### E. TEST-RETEST IN DYNAMIC RECONSTRUCTIONS

Two participants also underwent retest $^{18}$F-MK-6240 PET scans within 30 days of the baseline/Test PET scans to evaluate the test-retest reliability of the tau outcome measures.

The relative changes (RC%) between the two scans are summarized in Table S1. Reconstruction with COMBRA reduced the RC% for the eCN for most regions of interest. On the other hand, for the AD subject, the RC increased.

Here, MoCo had divergent effects. For low-motion eCN subjects, relative change (RC%) was reduced compared to both frame-to-frame (F2F) pipelines and no-motion correction (noMoCo) approaches. Conversely, high-motion AD subjects experienced increased RC% after applying COMBRA. This increase may reflect more accurate tracking of individual variability but warrants further investigation.

**Table S1** Test-retest relative change(%RC) for two MK subjects. Unlike the AD subject, the eCN subject shows smaller changes with the proposed motion compensation framework.

| | Precuneus | Entorhinal | InferiorTemporal | Amygdala | Parahippocampal | Hippocampus |
|---|---|---|---|---|---|---|
| **Test-Retest Subject A (eCN, age 65, Male, MMSE 29)** | | | | | | |
| **COMBRA** | -1.3% | 0.4% | -0.9% | 1.3% | -0.1% | 0.23% |
| **F2F** | 2.0% | 7.1% | -0.34% | -3.7% | 3.2% | 0.6% |
| **NoMoco** | 1.4% | 7.1% | -0.5% | -4.1% | 2.9% | -0.1% |
| **Test-Retest Subject B (AD, age 54, Male, MMSE 22)** | | | | | | |
| **COMBRA** | 13.7% | 10.6% | 13.0% | 11.4% | 17.7% | 19.2% |
| **F2F** | -7.1% | -4.2% | -5.0% | -2.3% | -14.4% | -8.9% |
| **NoMoco** | -2.4% | 1.3% | -0.4% | 1.8% | -5.9% | -3.4% |
